# An externally validated deep learning model for the accurate segmentation of the lumbar paravertebral muscles

**DOI:** 10.1101/2021.10.25.21265466

**Authors:** Frank Niemeyer, Annika Zanker, René Jonas, Youping Tao, Fabio Galbusera, Hans-Joachim Wilke

**Author notes:** Corresponding author: Fabio Galbusera, PhD; IRCCS Istituto Ortopedico Galeazzi, via Galeazzi 4, 20161 Milan, Italy.

## Abstract

**Purpos:** Imaging studies about the relevance of muscles in spinal disorders, and sarcopenia in general, require the segmentation of the muscles in the images which is very labour-intensive if performed manually and poses a practical limit to the number of investigated subjects. This study aimed at developing a deep learning-based tool able to fully automatically perform an accurate segmentation of the lumbar muscles in axial MRI scans, and at validating the new tool on an external dataset.

**Methods:** A set of 60 axial MRI images of the lumbar spine was retrospectively collected from a clinical database. Psoas major, quadratus lumborum, erector spinae, and multifidus were manually segmented in all available slices. The dataset was used to train and validate a deep neural network able to segment muscles automatically. Subsequently, the network was externally validated on images purposely acquired from 22 healthy volunteers.

**Results:** The Jaccard index for the individual muscles calculated for the 22 subjects of the external validation set ranged between 0.862 and 0.935, demonstrating a generally excellent performance of the network. Cross-sectional area and fat fraction of the muscles were in agreement with published data. *Conclusions*. The externally validated deep neural network was able to perform the segmentation of the paravertebral muscles in axial MRI scans in an accurate and fully automated manner, and is therefore a suitable tool to perform large-scale studies in the field of spinal disorders and sarcopenia, overcoming the limitations of non-automated methods.

## Introduction

The scientific study of sarcopenia gained high attention in recent years and is nowadays one of the most discussed issues in the field of the health of ageing subjects. According to the European Working Group on Sarcopenia in Older People, sarcopenia is defined as a “syndrome characterized by progressive and generalized loss of skeletal muscle mass and strength” [1]. Sarcopenia is a multifactorial condition associated with age-related reduction of physical activity, protein intake, anabolic hormonal activity, and vitamin D levels, as well as to a pro-inflammatory status due to increased intracellular oxidative stress [2]. The decrease of hormonal levels, especially testosterone, estrogen, and growth hormone, has been found to be associated with muscle wasting [3].

Several studies associated sarcopenia with spinal disorders. Toyoda and colleagues demonstrated that sarcopenia is correlated with back muscle strength and in turn with the presence of spinal degeneration [4]. The research highlighted that the association may be seen in two distinct ways: sarcopenia could induce spinal disorders, pain and spine-related disability, but also spinal disorders may cause muscle waste and increased fatty infiltration, probably due to the reduced physical activity associated with pain and fear-avoidance behavior. Indeed, sarcopenic patients were reported to show anxiety and catastrophizing signs more commonly than the age-matched population [5], initiating a vicious circle in which reduced physical activity due to psychological issues further promotes muscle waste. A correlation between sarcopenia and degenerative scoliosis, possibly involving reduced bone mineral density, has also been shown [6].

In general, the study of sarcopenia as a possible risk factor for spinal disorders, a consequence of it or its role as a risk factor enhancing symptoms and disability in patients suffering from spinal disorders is at its infancy, since most studies are very recent and have been conducted on relatively small patient cohorts. While the importance of sarcopenia has been undoubtedly recognized, large-scale studies quantifying it and investigating it in detail are still lacking.

Several imaging modalities, namely dual-energy X-ray absorptiometry (DXA), computed tomography (CT), magnetic resonance (MR), and ultrasound (US), have been used to investigate sarcopenia as well as for diagnostic purposes with the aim of quantifying muscle mass. MRI is recently gaining a wide interest since it allows measuring the muscle mass and the amount of fat infiltration inside muscle tissue, as well as additional information regarding muscular oedema, fibrous infiltration, fiber contractility, and elasticity [7], without any exposure to ionizing radiation. Fat and water discrimination can be done using multiecho gradient-echo sequences like the Dixon technique, which allows obtaining a quantitative assessment of intramuscular fat [8]. However, it should be noted that the use of MRI to study and diagnose sarcopenia requires the segmentation of the images, i.e. the identification of the contours of the muscle mass or of the individual muscles in each slice. Although semi-automated and fully automated methods to segment MRI scans have been presented [9–12], in most clinical papers about sarcopenia this step has been conducted by human operators since such methods are not publicly available and their performance is generally lower than that of expert operators, not generalizing well to pathological cases [11]. Besides, manual segmentation requires a substantial amount of work and practically poses a limit to the sample size, since processing high numbers of subjects such as several thousands requires a time not compatible with the length of most research projects.

The aim of this study is to develop and validate an accurate automated tool to perform the segmentation of axial MRI scans of the paravertebral muscles in the lumbar region, to be used for the assessment and diagnosis of sarcopenia as well as of the condition of the lumbar muscles in general. The automated method shall allow processing large amounts of data, thus allowing the imaging investigation of sarcopenia and its association with spinal disorders in large-scale populations. The secondary aim of the work is to externally validate the new algorithm in a population of 22 healthy volunteers, in order to quantitatively check its ability to deal with a wide scenario of ages and body sizes.

## Materials and methods

### Training data

A set of 60 axial images of the lumbar spine was retrospectively collected from the imaging database of University Hospital Ulm (Ulm, Germany). The retrospective data collection has been approved by the ethics committee of Ulm University (approval nr. 50/20). All slices relative to the L1-L5 region were manually segmented by means of ITK-SNAP free software (http://www.itksnap.org) by a single operator, using a published method as a reference [13]. The following muscles were segmented, depicting the left and right sides as separate entities: psoas major, quadratus lumborum, erector spinae, multifidus. The segmented images were randomly split into training and internal validation sets based on an 80-20% ratio.

### Network architecture

The architecture of the deep artificial neural network used for the segmentation of the muscles was derived from the two-dimensional U-Net [14], with several modifications: (1) Leaky ReLU (α = 0.01) and softmax as activation functions instead of ReLU and sigmoid; (2) transposed convolutions for upsampling; (3) dropout (p = 0.03) added to the encoder; (4) one additional layer in both encoder and decoder (Fig. 1). The network was trained using the Adam optimizer, with a learning rate of 0.001 and a minibatch size of 16. The architecture and the hyperparameters were extensively optimized by checking the performance of the network on the internal validation set.

**Figure 1.**
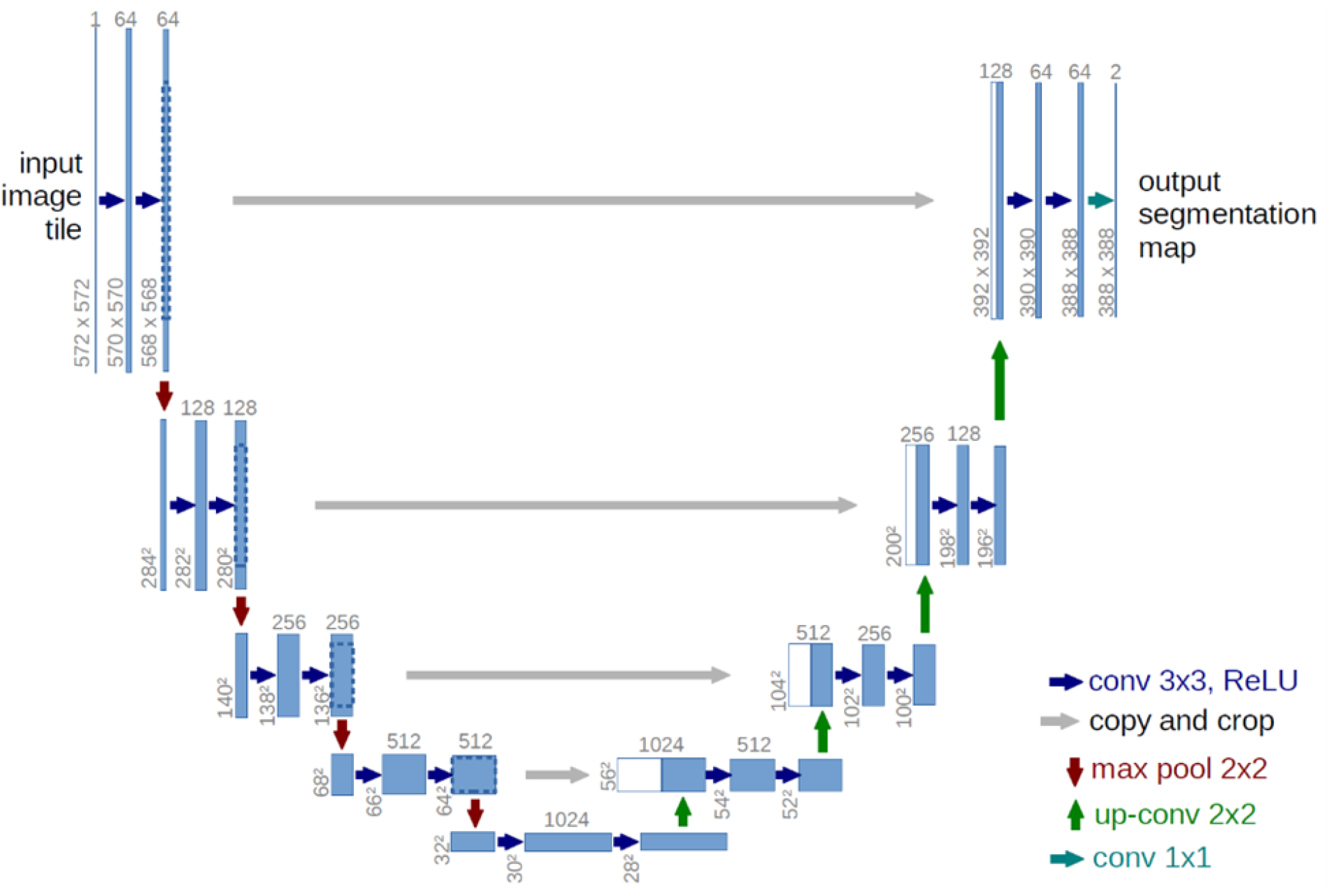
The architecture of the neural network derived from the two-dimensional U-Net employed in this study.

### Metrics

The “Intersection over Union”, in short *IoU* and also known as Jaccard similarity coefficient [15], was used as a metric to evaluate the quality of the segmentations generated by it. For a specific muscle *C*, the relative *IoU* can be calculated as follows:

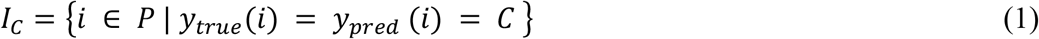

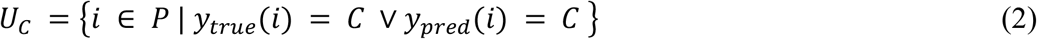

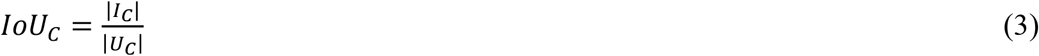

where *i* is a pixel belonging to the image *P*(intended as a set of pixels), *C*is the muscle of interest, *y*_*true*_(*i*) is the ground truth for the specific pixel *i, y*_*pred*_(*i*) is the value predicted by the neural network, *I*_*C*_ is the set of pixels corresponding to the muscle *C*in both the ground truth and the network output, *U*_*C*_ is the set of pixels corresponding to the muscle *C*in either the ground truth or the network output. The value of *IoU* is therefore 1 if the network output corresponds perfectly to the ground truth, and 0 if there is no overlap between the ground truth and the predictions.

For the sake of training the neural network, the sum of the *IoU* calculated among all segmented muscles was used as loss function:

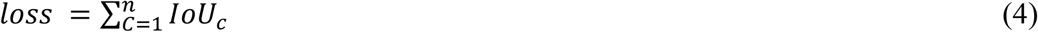

where *n* is the number of segmented muscles (8 in the present study).

### External validation

In order to test the performance of the segmentation tool on an external dataset, axial MRI scans of the whole lumbar spine were prospectively collected for 22 healthy volunteers, 10 males (mean age: 38.6 years (range 25-61), mean weight: 83.3 kg (68-120), mean height: 179.4 cm (170-185)) and 12 females (mean age: 33.5 years (range 22-57); mean weight: 57.5 kg (48-68); mean height: 166.3 cm (153-178)). The prospective image collection was approved by the ethics committee of Ulm University (approval nr. 255/20). The whole lumbar spine of all subjects was imaged with a 3T MRI system (MAGNETOM Skyra, Siemens Healthineers AG, Erlangen, Germany) by using standard imaging protocols. All images were manually segmented by the same operator who performed the segmentation of the training data using the same software tools and methods, as well as by the deep neural network. The manual segmentations were used as the reference for the quantitative assessment of the performance of the automated tool, which was conducted by calculating the *IoU* of the individual paravertebral muscles for all slices.

The segmented images generated by the neural network were used to calculate the cross-sectional area of the individual paravertebral muscles for the 22 volunteers, exploiting the information about pixel size and slice spacing included in the DICOM files (attributes (0028, 0030) and (0018, 0088) respectively). Furthermore, the fat fraction inside each muscle was determined by attributing each voxel to either fat or lean tissue, by using a simple Otsu binary thresholding implemented in the scikit-image Python library for image processing (https://scikit-image.org/).

## Results

The neural network provided excellent outputs from a qualitative point of view (Fig. 2). A three-dimensional reconstruction of the shape of the paravertebral muscles showed realistic anatomies from a perceptual point of view, with smooth surfaces and limited artefacts (Fig. 3). The median values of the *IoU* for the individual muscles calculated during the external validation ranged between 0.862 and 0.935 (Table 1), demonstrating a generally excellent agreement between the ground truth and the outputs of the neural network. The psoas major and the erector spinae showed a generally higher performance with respect to the quadratus lumborum and the multifidus (Fig. 4), suggesting that a higher cross-sectional area was associated with a higher *IoU*. Nevertheless, the lower quartile of the statistical distribution of the *IoU* was in all cases higher than 0.75, indicating a high quality of the output in the vast majority of the processed slices.

**Table 1.** Median values of the *IoU* for the individual paravertebral muscles calculated for the 22 volunteers recruited for the external validation.

|  | psoas major |  | quadratus lumborum |  | erector spinae |  | multifidus |  |
| --- | --- | --- | --- | --- | --- | --- | --- | --- |
|  | *left* | *right* | *left* | *right* | *left* | *right* | *left* | *right* |
| *IoU* | 0.930 | 0.935 | 0.891 | 0.892 | 0.929 | 0.923 | 0.868 | 0.862 |

**Figure 2.**
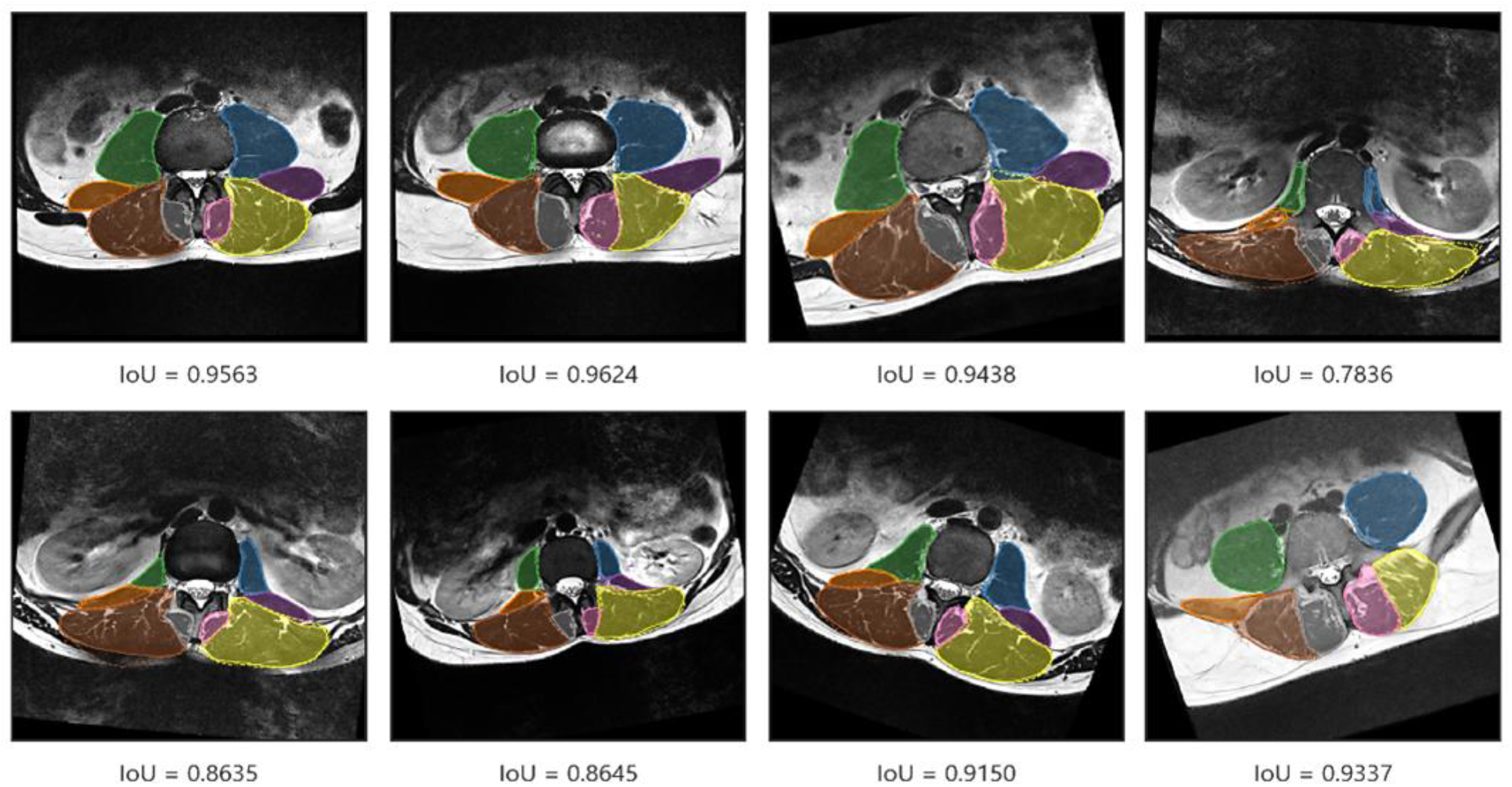
Segmentations of the paravertebral muscles for randomly selected slices of eight representative subjects recruited for the external validation. The average *IoU* for the eight muscles is also shown. Green/blue: psoas major; orange/purple: quadratus lumborum; brown/yellow: erector spinae; gray/pink: multifidus.

**Figure 3.**
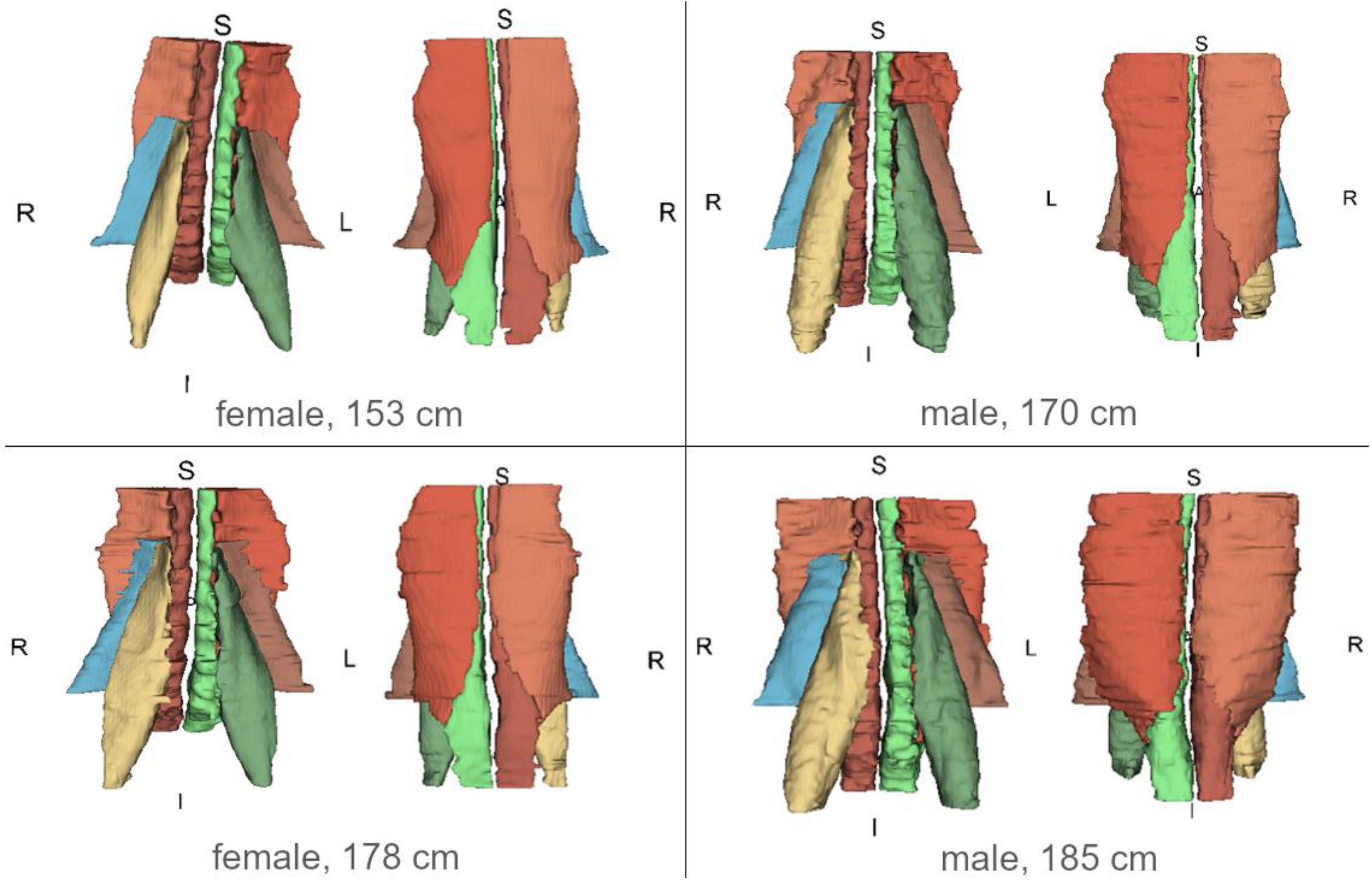
Three-dimensional reconstructions of the segmented paravertebral muscle for four representative volunteers. R: right; L: left; S: superior; I: inferior.

**Figure 4.**
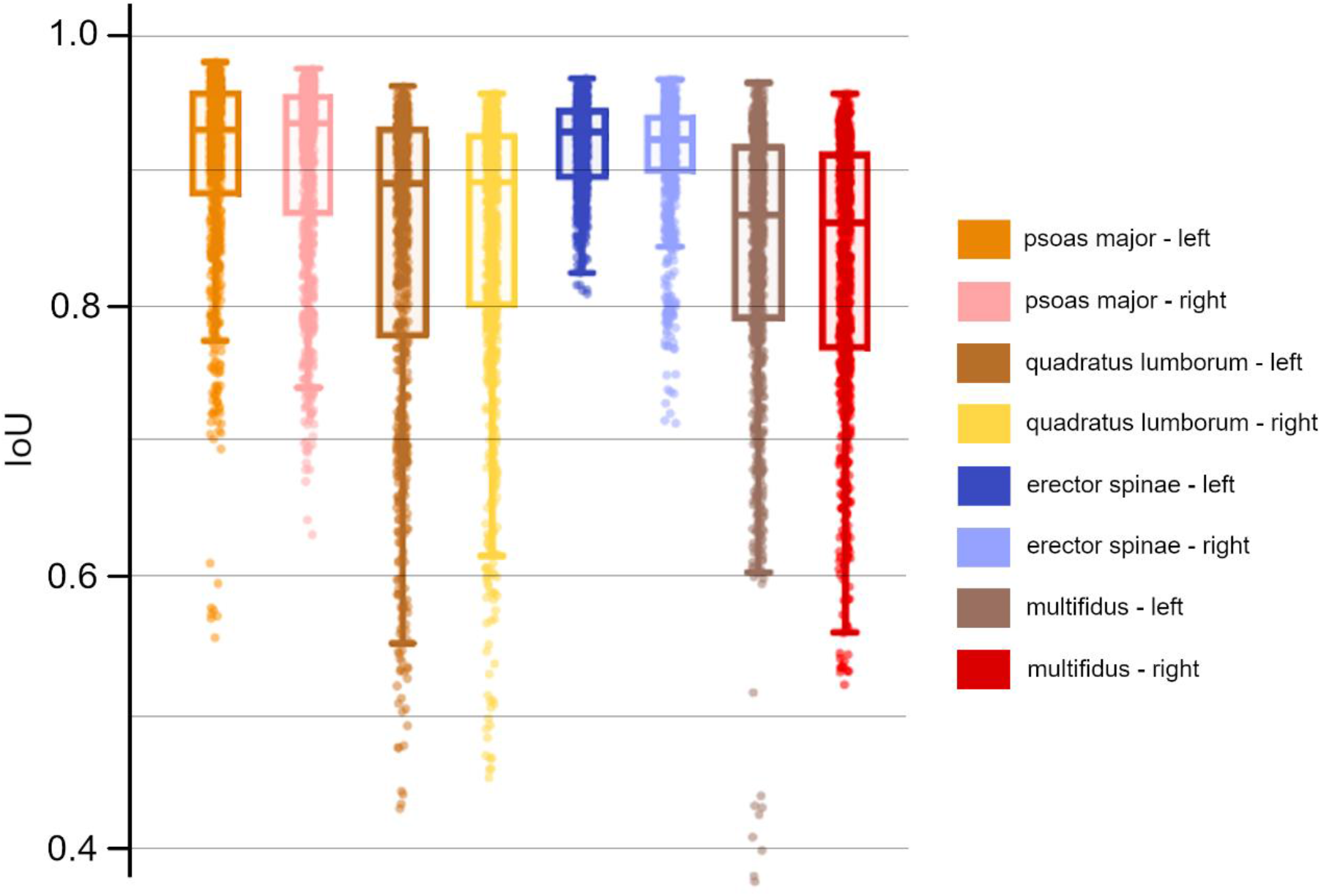
Boxplots and jittered scatter plots of the *IoU* calculated for the individual muscles in all slices acquired for the 22 subjects recruited for the external validation.

Among the 22 subjects included in the external validation, the male subjects showed generally higher cross-sectional areas of all muscles with respect to females (Fig. 5). The erector spinae was the larger muscle in all volunteers, followed by the psoas major in males while in females the size of the latter muscle generally overlapped those of the quadratus lumborum and multifidus. The fat fraction was larger in females than in males (Fig. 6) and generally increased with age. A non-negligible variability of the fat fraction was found among the individual muscles, with the multifidus and the erector spinae generally showing higher fatty infiltration with respect to psoas major and quadratus lumborum.

**Figure 5.**
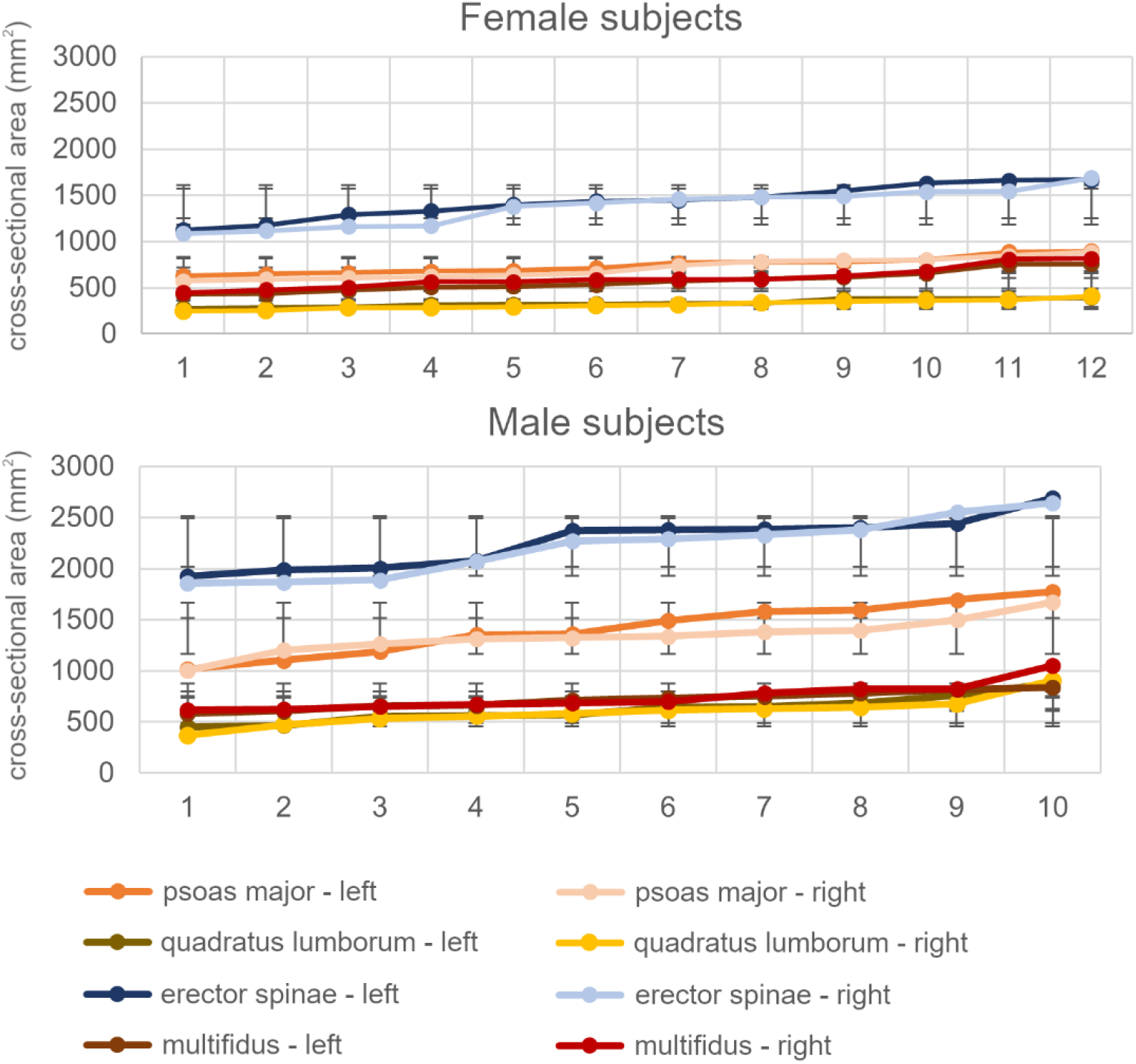
Cross-sectional areas (mean value and standard deviation, in mm^2^) of the eight paravertebral muscles for the 22 subjects (top: females, bottom: males) in ascending order.

**Figure 6.**
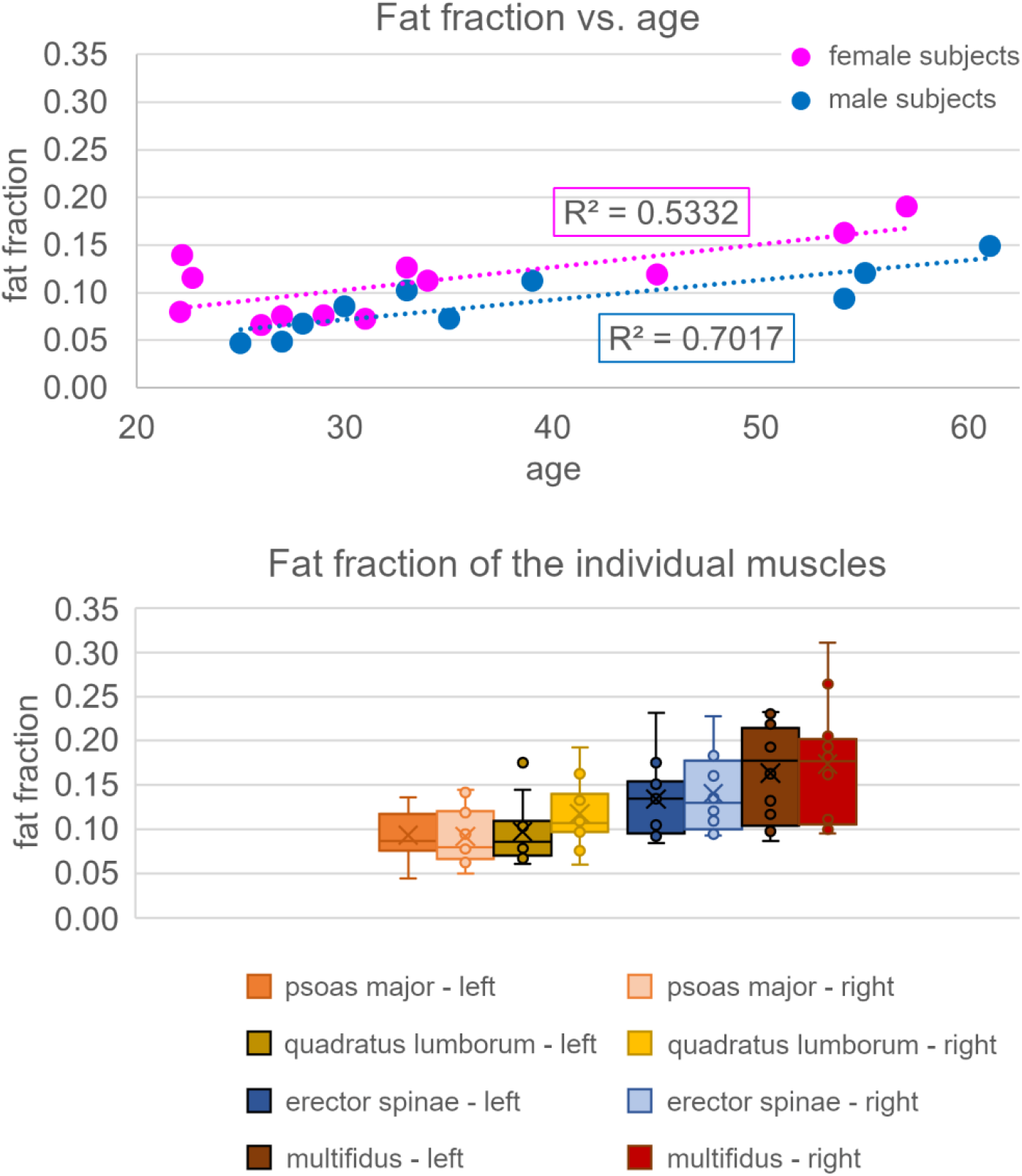
Fat fraction in the paravertebral muscles depending on age and stratified by sex (top) and for the individual muscles (bottom) for the 22 subjects.

## Discussion

In this paper, we presented a novel tool based on artificial intelligence to perform the automatic segmentation of the paravertebral muscles in axial MRI scans. The tool demonstrated excellent performance, potentially enabling its use for processing large amounts of data in retrospective population studies about sarcopenia and its relation with spinal disorders. Since manual segmentation has always constituted a bottleneck for imaging studies about muscles due to its high requirements in terms of manual labour, existing works have been generally limited to dozens or hundreds of cases (e.g. [16, 17], with large populations consisting of several thousand subjects being practically not manageable. An accurate segmentation tool that does not require any human intervention can therefore act as the key enabling technology for large-scale studies. Sarcopenia, in particular in terms of reduced mass and quality of the back muscles, is associated with higher disability and lower quality of life in patients suffering from spinal disorders with respect to non-sarcopenic patients [18]. Sarcopenic patients subjected to spine surgery have a slower and sometimes less complete recovery, more severe postoperative symptoms and lower satisfaction, whereas the correlation between sarcopenia and increased risk of postoperative complications could not be proved [19]. In general, despite the literature indicating that sarcopenia is a highly relevant risk factor for patients suffering from spinal disorders in terms of worse outcomes, relatively little research has been performed in the field, and the availability of fully automated imaging segmentation tools can definitely play a role in future studies addressing this need.

Other automated solutions based on neural networks aimed at segmenting the lumbar paravertebral muscles have been previously presented. Li et al. [12] developed a U-Net based network to perform the segmentation of multifidus and erector spinae, obtaining average Dice similarity coefficients of 0.949 and 0.913 respectively, generally in line with the performance of the current model while taking into account that the metrics used in the present and in the literature study are not directly comparable. Zhang et al. developed another U-Net based model and achieved similar performances [20]. However, it should be noted that in both studies psoas major and quadratus lumborum were not segmented, and that an internal validation on a test set randomly selected from the original database rather than an external validation of a distinct dataset was performed. Xia and colleagues [11] tested several network architectures, including the U-Net as well as novel solutions, to segment psoas major, erector spinae, and multifidus, obtaining excellent Dice similarity scores between 0.913 and 0.95 for the best-performing neural network. Other studies achieved relatively lower performances [21], or focused on different imaging modalities or fields of view [22, 23]. In general, it can be concluded that our model achieved state-of-the-art performance and is the only one that has been externally validated on a purposely created dataset so far.

The results extracted from the population recruited for the external validation, i.e. the cross-sectional areas of the individual paravertebral muscles and their fat fraction, were in good agreement with existing research, further consolidating the validity of the novel model. Cooper et al. measured cross-sectional areas very similar to those in the present study for the paraspinal muscles (erector spinae and multifidus) as well as for the psoas major as well as the same gender-based differences, including the same disproportion between the size of the psoas in males and females with respect to the other muscles [24]. Fat fractions were also in line with previous observations [16, 25].

The study suffers from some limitations. First, the manual segmentations of the training set and for the external validation were performed only once by a single operator, and a quantitative assessment of the reliability and quality of the ground truth data could therefore not be performed. Besides, a single network architecture was employed as a basis for the development and optimization of the model; while U-Net is largely considered as a state-of-the-art solution for the segmentation of medical images [26], other solutions such as for example generative adversarial networks may be worthy of investigation. Finally, the subjects recruited for the external validation were in a relatively low number and, while they covered a rather wide range of age and body sizes, were all asymptomatic. Therefore, the performance of the tool was not validated for severe pathological cases, which can be nevertheless highly relevant from a clinical point of view.

In conclusion, the tool here presented was able to perform the automated segmentation of the paravertebral muscles in axial MRI scans in an accurate manner in an external validation set of patients, and is therefore a suitable tool to perform future large-scale studies aimed at analyzing the size and quality of the muscles of the lumbar region at the population level.

## Data Availability

All data produced in the present study are available upon reasonable request to the authors.

